# White matter microstructural changes in adult-onset idiopathic focal cervical dystonia using ultra-strong diffusion gradient MRI

**DOI:** 10.1101/2024.02.07.24302448

**Authors:** CL MacIver, DK Jones, K Green, K Szewczyk-krolikowski, A Doring, CMW Tax, KJ Peall

## Abstract

**Background and Objectives:** Adult-onset idiopathic focal cervical dystonia (AOIFCD) involves loss of co-ordinated contraction of the cervical musculature, resulting in pain, impaired function and in some individuals, an associated head tremor. Existing neuroimaging studies have implicated key motor networks. However, measures used to date lack specificity in detailing the underlying pathophysiological differences.

**Methods:** A cohort of individuals diagnosed with AOIFCD and an age- and sex-matched control group were prospectively recruited. All participants underwent MRI using structural and diffusion sequences with multiple b-values up to 30,000 s/mm2, coupled with motor and non-motor clinical phenotyping. Tractography was performed assessing whole tract median values, while tractometry was used for along tract analysis. Key white matter motor pathways were assessed initially using general measures (DTI/DKI: FA-fractional anisotropy; MD-mean diffusivity; MK-mean kurtosis; AK-axial kurtosis; RK-radial kurtosis) with subsequent microstructural white matter modelling approaches (NODDI: ODI-orientation distribution index, NDI-neurite density index, FWF-free water fraction; and standard model: *f*-intra-axonal signal fraction, D_a_-intra axonal diffusivity, D_epar_-extra axonal parallel diffusivity, D_eperp_-extra axonal perpendicular diffusivity, *p*_2_-orientation coherence) and unconstrained high b-value zero-order spherical harmonic signal (R0, related to intra-axonal signal) to assess differences within these tracts. Subgroup analyses were undertaken comparing those with and without associated head tremor to the control cohort.

**Results:** 50 AOIFCD and 30 healthy control participants underwent structural brain MRI, with 46 AOIFCD and 30 healthy controls included for analysis (33 without head tremor, 13 with head tremor). Significant differences were observed in the anterior thalamic radiations (lower mid tract FA, RK, *f* and *p*_2_ and higher ODI), thalamopremotor tracts (mid tract higher MK and lower NDI, and distal tract lower ODI and higher *f*) and striatopremotor tracts (proximal lower *f* and R0). These measures correlated with symptom severity across the spectrum with clinical measures, including psychiatric symptoms, sleep quality, pain and cognitive functioning.

**Discussion:** Overall, localised microstructural differences were identified within tracts linking the prefrontal cortex and premotor cortex with basal ganglia regions, suggesting microstructural aberrances of motor system modulatory pathways, particularly in relation to intra-axonal and fibre orientation dispersion measures.

## Introduction

Adult-onset idiopathic focal cervical dystonia (AOIFCD) is the most common form of adult-onset idiopathic dystonia. Cervical muscle over-activity and the loss of co-ordinated muscle contraction lead to abnormal posturing and pain, with a proportion of patients experiencing an associated head tremor. Pathophysiological understanding of AOIFCD is limited, however findings from human functional brain imaging studies of focal dystonia indicate reduced motor network based functional connectivity(1, 2) and disruption to inhibitory/excitatory neurotransmitter balance, including reduced thalamic GABA levels(3). With respect to structural imaging, grey matter volumetric differences have been most consistently demonstrated amongst task-specific dystonias with increased volumes in motor grey matter(4), and more variable findings observed in AOIFCD cohorts(5, 6). White matter analysis in AOIFCD has predominantly involved diffusion MRI, using relatively non-specific measures, coupled with heterogenous approaches and findings. Some have demonstrated localised higher, lower or unchanged diffusion tensor measures such as fractional anisotropy (FA) and mean diffusivity (MD) within motor pathways (5, 7–10).

By contrast, monogenic forms of dystonia have demonstrated more consistent findings, including lower FA in motor pathways connecting the cerebellum, basal ganglia structures and cortical motor grey matter, coupled with indications of more widespread white matter pathology (11–14). More recently, tremor specific imaging differences have been suggested, with higher cerebellar peduncular FA observed in dystonic tremor compared to other tremor syndromes (8, 9). These differences between disparate genotypes and phenotypes of dystonia may suggest that interpretations such as lower white matter tract integrity are not a unifying feature across the spectrum of dystonic disorders. Additionally, parameters such as FA based on diffusion tensor MRI, comprising most of the existing literature, are of a non-specific nature and have the potential to represent a range of underlying biological changes(15).

To date, few attempts have been undertaken to assess more microstructurally relevant parameters, including utilising modelling and other approaches which compartmentalise the intra- and extra-axonal signal with the aim to more specifically quantify tissue characteristics. One fixel-based analysis approach has described lower apparent fibre density in the region of the striatum(16) in AOIFCD, and regions of both higher and lower orientation dispersion using Neurite Orientation Dispersion and Density imaging (NODDI) in the superior cerebellar peduncles and anterior thalamic radiations in a mixed dystonia cohort(9). These approaches, however, have been undertaken using acquisitions which consist of only low to moderate b values (up to b=2000s/mm^2^), with potential for inaccuracy in modelled parameters stemming from the *a priori* fixing of values to address model degeneracy (17). To overcome some of these limitations, diffusion MRI approaches which make use of more advanced acquisitions, such as multiple b-value measurements that include higher b-values (>2000 s/mm^2^) or multiple b-tensor encoding schemes(18), have been exploited to optimise modelling approaches and negate the need for extensive *a priori* fixing of parameters (19).

Here, we use ultra-strong diffusion gradient MRI scanning in a deeply phenotyped AOIFCD cohort to explore white matter microstructural differences in pathways linking key motor brain hubs, and if present, anticipate accentuation of these differences through application of optimal acquisition and microstructural modelling approaches.

## Methods

### Recruitment

Participant recruitment was via the Welsh Movement Disorders Research Network (REC reference: 14/WA/0017 IRAS ID: 146495), with clinical phenotyping performed under the Global Myoclonus Dystonia Registry and Non-Motor Symptoms Study (REC reference: 18/WM/0031, IRAS project ID: 236219). Ethical approval for the brain imaging was obtained via the Cardiff University School of Medicine Research Ethics Committee (SMREC reference number 18/30). Study inclusion required a clinical diagnosis of AOIFCD, diagnosed by a neurologist with expertise in movement disorders, with exclusion criteria including existence of a contraindication to MR scanning, significant claustrophobia, inability to tolerate the scan, co-existence of other neurological diagnoses, a diagnosis of another form of dystonia.

### Clinical characterisation

During the assessment visit, a videotaped clinical examination was undertaken following a modified form of the Burke-Fahn-Marsden dystonia scale (BFMDRS) protocol that would also allow for scoring with both the BFMDRS and Toronto West Spasmodic Torticollis Rating Scale (TWSTRS). These were reviewed and scored independently by two neurologists with expertise in movement disorders.

Extensive non-motor phenotypic data was additionally acquired. The participants undertook a self-completed questionnaire assessing basic demographic information, psychiatric symptomatology (the modified Mini International Neuropsychiatric Interview score (MMS), Structured Clinical Interview for DSM-5 Personality Disorders, Beck’s Depression inventory, Health Anxiety Index and the Yale-Brown Obsessive Compulsive scale), sleep (Pittsburgh Sleep Quality Index (PSQI) (20), The Sleep Disorders Questionnaire (SDQ) (21) and The Epworth Sleepiness Scale), pain (typical pain time course, Chronic Pain Acceptance Questionnaire, Pain Catastrophising scale) and quality of life (Short Form 36 Health survey). Cognitive function was assessed face-to-face with the National Adult Reading Test (NART) used to determine IQ, and the Cambridge Neuropsychological Test Automated Battery (CANTAB, www.cantab.com) used to assess multiple cognitive domains (One Touch Stockings of Cambridge (executive function), Emotional Recognition task, Paired Associated Learning and Spatial Working Memory).

### MRI data acquisition

#### Scanning acquisition

All imaging was undertaken on a 3T Connectom scanner (Siemens Healthcare), with a 32-channel head coil and a scan time for white matter assessment of approximately 45 minutes. An initial structural scan was undertaken using a sagittal T1-weighted MPRAGE acquisition with the acquisition using a TR of 2300ms, TE 2ms, flip angle 9° and inversion time 857ms. The field of view was 192mm x 256mm x 256mm, with a voxel size of 1mm x 1mm x 1mm. Diffusion MRI data were acquired for the whole brain with multiple b-values (b=0s/mm^2^ (19 directions), b=200 s/mm^2^ (20 directions), b=500 s/mm^2^ (20 directions), b=1200 s/mm^2^ (30 directions), b=2400 s/mm^2^ (61 directions), b=4000 s/mm^2^ (61 directions), b=6000 s/mm^2^ (61 directions), b=30000 s/mm^2^ (120 directions). Gradient sampling vectors were distributed across the entire unit sphere. The acquisition used a TR 4000ms and a TE of 75ms, Δ 30ms and δ 15ms, flip angle 90°. The field of view was 110mm x 110mm x 66mm, with a voxel size of 2mm x 2mm x 2mm. No partial Fourier was used. An additional b=0 image was acquired with a reversed phase encoding direction.

### Preprocessing

The overall preprocessing and analysis approach is summarised in Figure 1A. The T1-weighted acquisition underwent on-scanner distortion correction, while the diffusion acquisition underwent brain extraction, denoising (MP-PCA based approach)(22), outlier detection (e.g., due to signal dropout from motion) (23), signal drift correction, and correction for Gibbs ringing(24), eddy currents, susceptibility distortion and subject motion (using FSL (version 6.0.5) eddy)(25) and gradient non-linearities(26, 27). The FSL eddy step included ‘repol’ (where outlier slices were removed and replaced using estimates) and ‘slice-to-vol’ motion correction for the participants with significant between-slice motion (where individual slices moved by motion were realigned to the overall volume).

**Figure 1:**
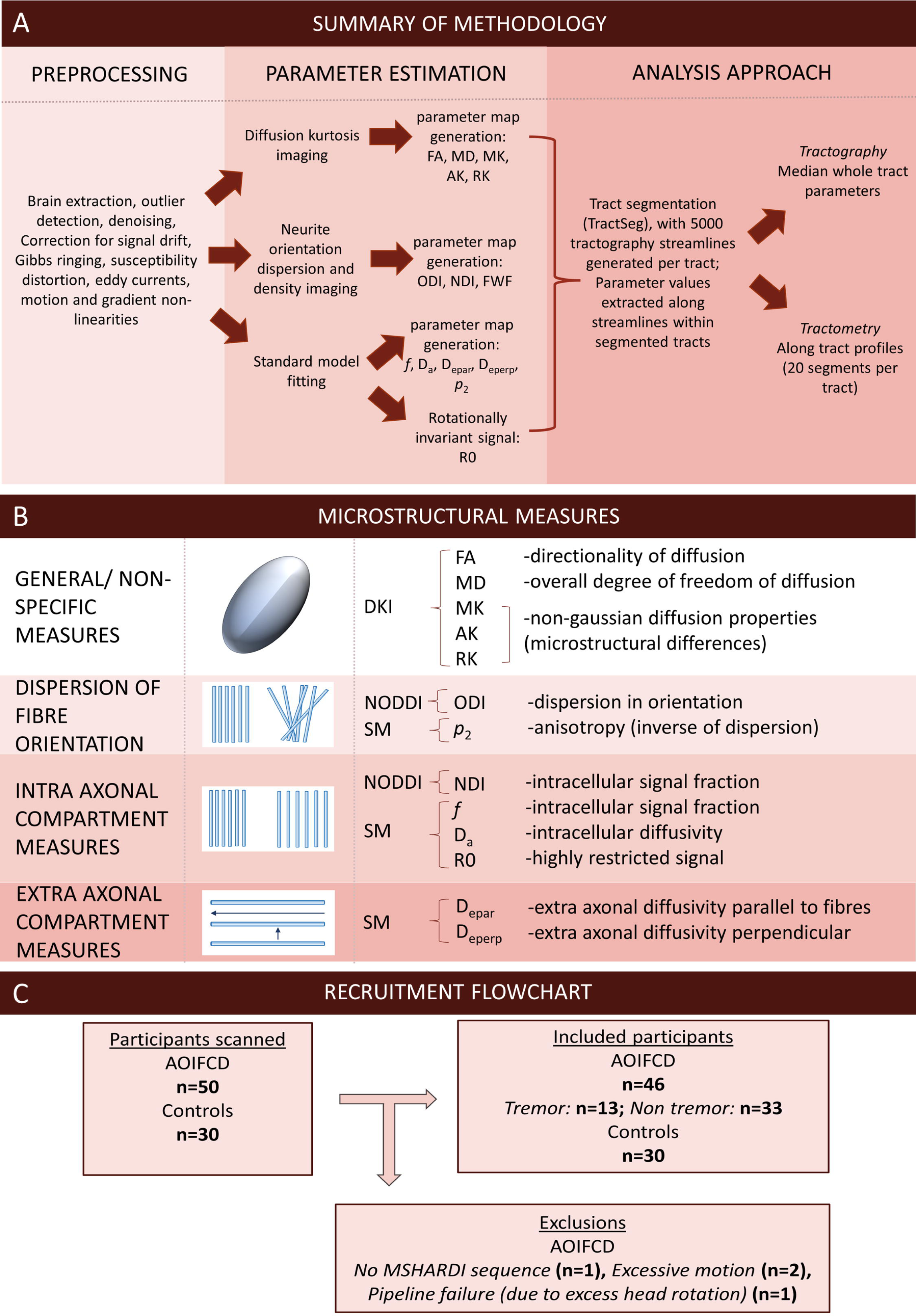
A) Overview of analysis methodology. B) Schematic of measured parameters. C) Recruitment flowchart. FA-fractional anisotropy; MD- mean diffusivity; MK- mean kurtosis; AK- axial kurtosis; RK- radial kurtosis; ODI- orientation distribution index; NDI- neurite density index; FWF- free water fraction; *f*- intraneurite signal fraction; D_a_- intra axonal diffusivity; D_epar_- extra axonal parallel diffusivity; D_eperp_- extra axonal perpendicular diffusivity; *p*_2_- orientation coherence; R0- rotationally invariant signal for order=0, b=6000 s/mm2; DKI- diffusion kurtosis imaging; NODDI- neurite orientation dispersion and density index; SM- standard model.

### Diffusion parameter estimates

1. Diffusion tensor imaging (DTI) and diffusion kurtosis imaging (DKI): Diffusion tensor (FA, MD) and diffusion kurtosis (MK- mean kurtosis, AK-axial kurtosis, RK-radial kurtosis) parameters were estimated using a constrained weighted least squares fitting approach with weight reduction for outlier measurement (27, 28).
2. Microstructural modelling: a) NODDI: estimation of parameter maps for orientation dispersion index (ODI), neurite density index (NDI), and free water fraction (FWF)(29), b) Standard model of white matter: a more general white matter model was fit using fewer constraints. This approach involved a least squares estimation of rotationally invariant spherical harmonics of the diffusion signal up to L=4, using the acquired b-shells up to b=6000 s/mm^2^, with Rician bias correction included. From this, the Standard Model parameters were estimated with data driven machine learning regression(30), applying a three-compartment model (Intra-axonal signal, extra axonal signal and free water). Parameter maps were estimated for *f* (intra-axonal signal fraction), D_a_ (intra-axonal diffusivity), D_epar_ (extra-axonal parallel diffusivity) and D_eperp_ (extra-axonal perpendicular diffusivity) and *p*_2_ (measure for orientation coherence). In addition to the modelled parameters, the raw rotationally invariant signal for L=0, b=6000 s/mm^2^ (R0) was assessed without addition of constraints or model fitting, considered reflective of intra-axonal signal(31).

Across the modelling approaches, parameters aimed to represent orientational dispersion of axon bundles (NODDI: ODI- the degree of dispersion of axons; SM: *p*_2_- the degree of orientation coherence, therefore inversely related to the ODI); measures relating to aspects of intra-axonal signal generally taken to be reflective of axonal density (NODDI: NDI- the proportion of signal attributable to the intra axonal compartment; SM: *f*- the proportion of signal attributable to the intra-axonal compartment, D_a_- the degree of diffusivity in the intra axonal compartment; R0-: rotationally invariant spherical harmonics of the signal (at L=0, b=6000) hypothesised to correspond to intra axonal signal); and measures relating to the extra axonal compartment (SM: D_epar_- the degree of diffusivity in the extra axonal space parallel to the axonal compartment, D_eperp_- the degree of diffusivity in the extra axonal space perpendicular to the axonal compartment)(Figure 1B).

### Tract segmentation

White matter tracts of interest were segmented using Tractseg(32), replicating our previous approach in an undifferentiated dystonia cohort(9): middle cerebellar peduncle, bilateral inferior cerebellar peduncles, superior cerebellar peduncles, frontopontine tracts, corticospinal tracts, anterior thalamic radiations, superior thalamic radiations, thalamoprefrontal tracts, thalamopremotor tracts, thalamoprecentral tracts, thalamopostcentral tracts, striatopremotor, striatoprecentral tracts, striatopostcentral tracts and optic radiations (used as a non-motor comparator tract). Multi-shell multi-tissue constrained spherical deconvolution was applied to the preprocessed diffusion data, with peak-extraction (maximum of three peaks/voxel). A fully connected convolutional neural network then created a tract probability image for each orientation and tract. Start and end regions were segmented, with fibre orientation maps calculated within each segmented tract(33).

### Analysis

Probabilistic tractography(34) with 5000 streamlines was performed for each tract. Median values for each parameter were extracted within segmented tracts for the tractography analysis. Along-tract profiling (tractometry) was performed for each parameter(35, 36) given the implication from previous work that white matter differences are localised rather than tract-wide(9). Tracts were split lengthwise into 20 equal segments, identifying the centroid of all streamlines within each segment. The end two segments were excluded, with the median parameter value across streamlines calculated for each of the 18 remaining segments(36). Tractometry was not performed for the thalamoprefrontal, thalamoprecentral, striatoprecentral, thalamopostcentral or striatopostcentral tracts as their geometry doesn’t allow for consistent segmentation.

### Excluded data

Following MR scanning, participants were excluded if there was significant motion artifact evident on the preprocessed diffusion acquisitions, assessed using FSL EddyQC and via manual inspection.

### Statistical Analysis

Analysis was undertaken using RStudio version 0.99.892. Participant demographic and clinical phenotypic data was summarised using appropriate descriptive statistics dependent upon data distribution. Regression models were applied comparing disease status (independent variable) for each parameter (dependent variable), with age, gender and handedness included as covariates.

Additional analyses were performed for those with and without associated tremor, with each group being compared to the control cohort. Bonferroni correction for multiple comparisons was undertaken for parameters, with differences reported only if significant following correction for multiple comparisons, unless otherwise stated. Correlation analyses were used to examine for association between the clinical phenotypic traits and the significant diffusion parameters using Pearson’s correlation coefficients.

## Results

Fifty individuals diagnosed with AOIFCD and 30 age- and sex-matched control participants were recruited (p=0.919 and p=0.912 respectively). Four participants from the AOIFCD cohort were excluded due to incomplete imaging acquisition (n=1), excess movement artefact (n=2) and failure of the analytical pipeline (n=1) (Figure 1C). The median age at examination of the AOIFCD cohort was 55.5 years (range 30-74 years), and 57 years (range 32-72 years) in the control cohort. The median age at onset of dystonia onset was 43.5 years (range 30-67 years) and median time since dystonia diagnosis 12.5 years (range 1-33 years). Demographic characteristics are summarised in Table 1, with no significant differences observed except for a lower median IQ score within the tremor cohort (median=111.4) compared to controls (118.9) (p=8.14×10-^3^). An overview of the non-motor phenotypic characteristics of the cohort is summarised in Supplementary Table 1. A significantly higher burden of these symptoms was observed across all non-motor domains in the AOIFCD cohort, including overall psychiatric symptomatology (p=2.49×10^-4^), depression (p=0.0199), sleep disturbance (p=0.004 (Pittsburgh Sleep Quality Index), p=5.78×10^-5^ (Sleep Disorders Questionnaire)), pain (p=0.004), spatial working memory (p=0.003) and overall quality of life (p=2.31×10^-4^).

**Table 1:**
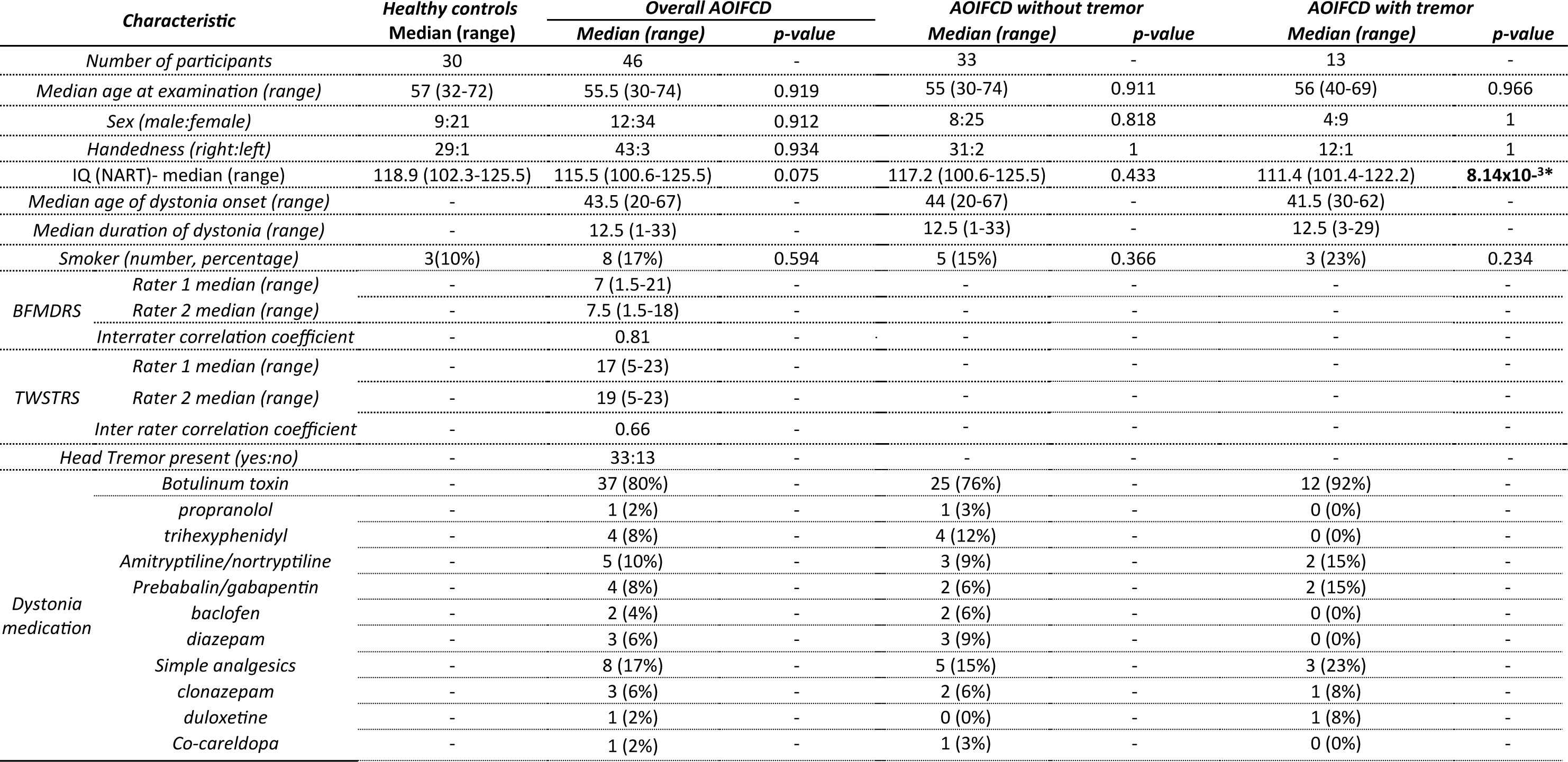
Summary of clinical characteristics of participants across cohorts. NART- National Adult Reading Test; BFMDRS- Burke Fahn Marsden Dystonia Rating Scale; TWSTR- Toronto West Spasmodic Torticollis Rating Scale. Significant differences are indicated with *.

### Tractography

Tractography analysis demonstrated no significant differences in median values between the overall AOIFCD cohort, tremor, or non-tremor subgroups, compared to controls for any of the measured parameters including DTI/DKI, NODDI, standard model, or rotationally invariant spherical harmonics (Supplementary Figures 1, 2, and 3).

### Tractometry

#### Overall dystonia cohorts vs. controls

Localised significant differences were observed in the anterior thalamic radiations (Figure 2), thalamopremotor tracts (Figure 3) and striatopremotor tract (Figure 4). In the anterior thalamic radiations FA values were significantly lower on both right (p=2.68×10^-3^) and left side (p=3.07×10^-3^) coupled with corresponding lower RK (p=1.42×10^-4^) and higher ODI NODDI values (p=2.22×10^-3^) on the left. The Standard Model analysis showed lower associated *p*_2_ (p=1.64×10^-3^) and *f* (p=2.78×10^-3^), again on the left side. The thalamopremotor tracts revealed right sided mid and distal-tract differences, with higher mid-tract MK (p=7.56×10^-4^) and associated lower NDI (p=2.1×10^-3^), with distal-tract higher ODI (p=3.1×10^-3^) and lower f (p=2.3×10^-3^). Changes in the striatopremotor tract were observed in the proximal portion of the right tract with lower *f* (p=1.06×10^-3^) (standard model) and corresponding lower R0 (p=2×10^-3^) (unconstrained rotational invariant signal measure).

**Figure 2:**
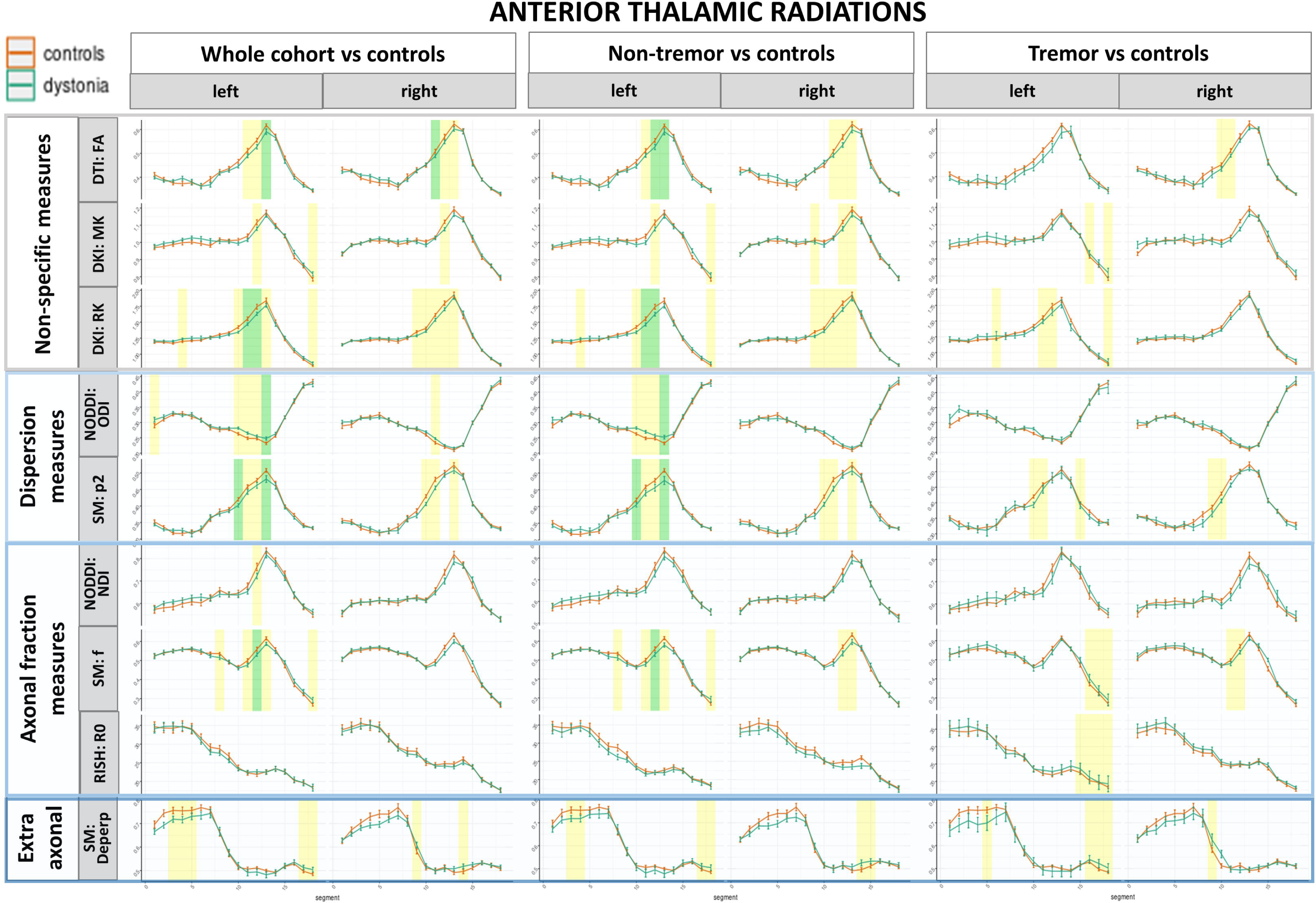
Along tract profiles for the anterior thalamic radiations, showing only parameters which showed significant differences in any analyses, with median tract value and standard error for each cohort. Green shaded regions represent those significantly different following multiple comparison correction, and yellow shaded regions are significant prior to correction. DTI- diffusion tensor imaging; FA- fractional anisotropy; DKI- diffusion kurtosis imaging; MK- mean kurtosis; RK- radial kurtosis; NODDI- neurite orientation dispersion and density imaging; SM- standard model; ODI- orientation dispersion index; *p*_2_- orientational coherence; NDI- neurite density index; *f*- intra neurite signal fraction; RISH- rotationally invariant spherical harmonics; R0- rotationally invariant signal for order=0, b=6000 s/mm^2^; D_eperp_- extra axonal perpendicular diffusivity.

**Figure 3:**
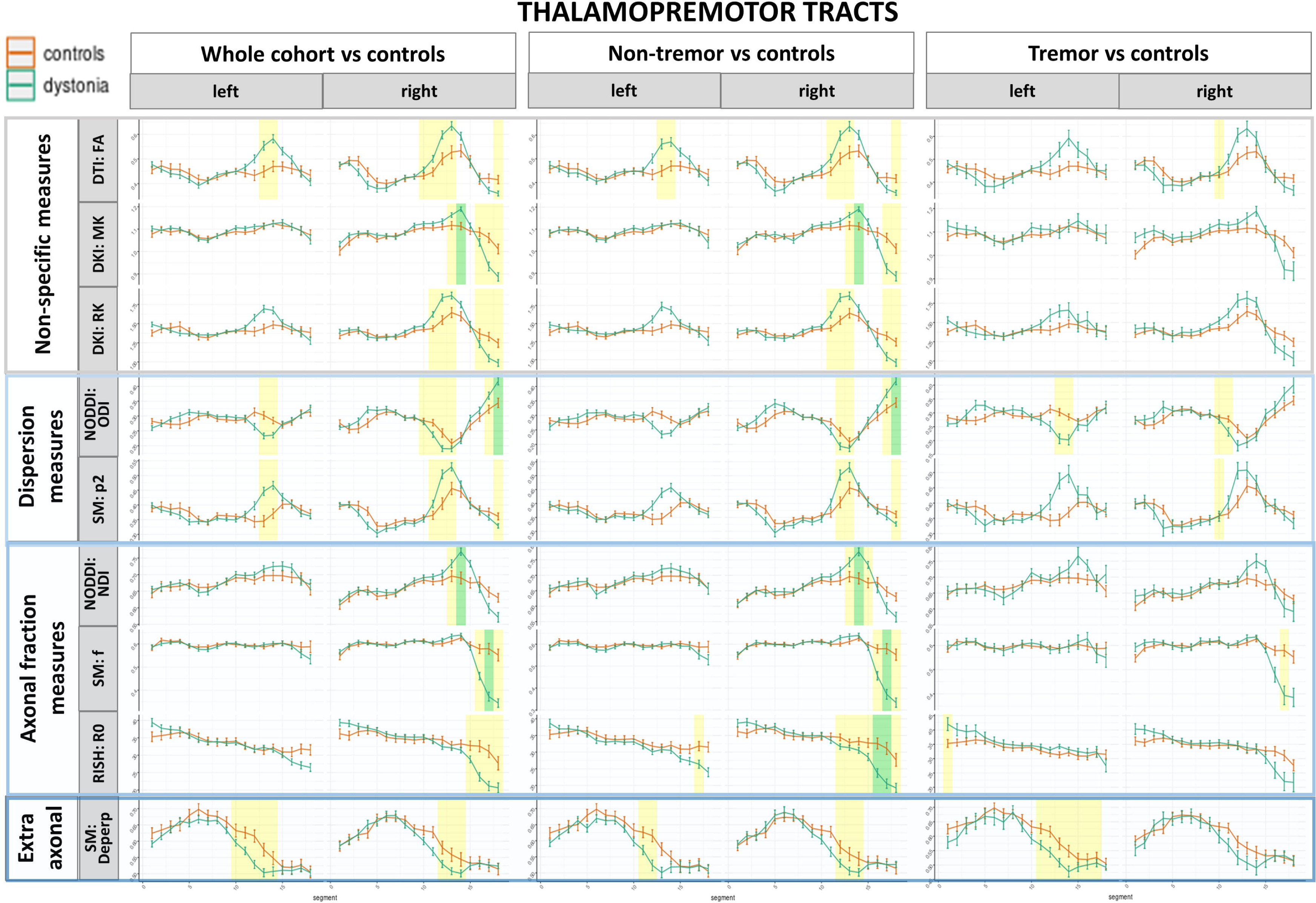
Along tract profiles for the thalamopremotor tracts, showing only parameters which showed significant differences in any analyses, with median tract value and standard error for each cohort. Green shaded regions represent those significantly different following multiple comparison correction, and yellow shaded regions are significant prior to correction. DTI- diffusion tensor imaging; FA- fractional anisotropy; DKI- diffusion kurtosis imaging; MK- mean kurtosis; RK- radial kurtosis; NODDI- neurite orientation dispersion and density imaging; SM- standard model; ODI- orientation dispersion index; *p*_2_- orientational coherence; NDI- neurite density index; *f*- intra neurite signal fraction; RISH- rotationally invariant spherical harmonics; R0- rotationally invariant signal for order=0, b=6000 s/mm^2^; D_eperp_- extra axonal perpendicular diffusivity.

**Figure 4:**
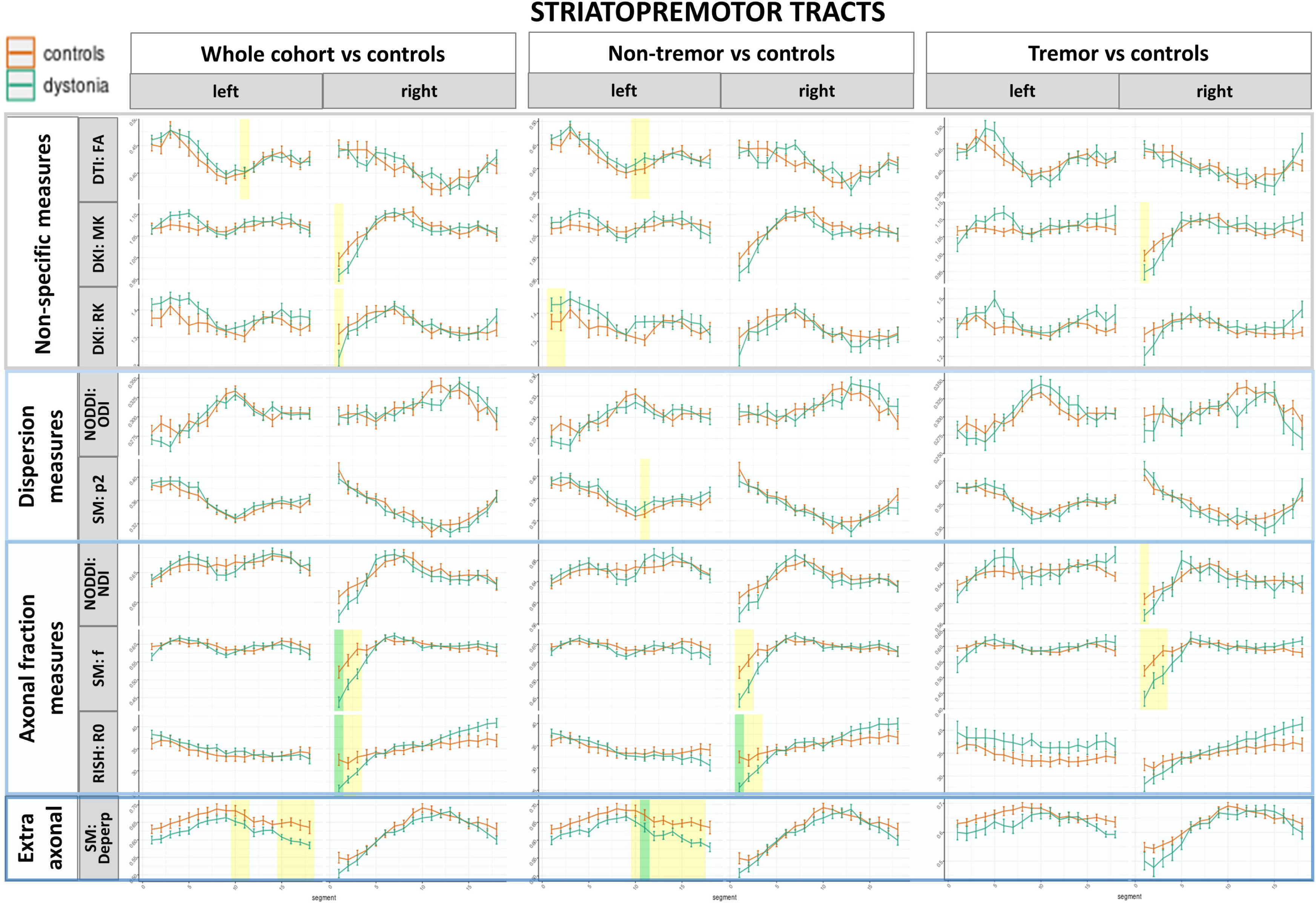
Along tract profiles for the striatopremotor tracts, showing only parameters which showed significant differences in any analyses, with median tract value and standard error for each cohort. Green shaded regions represent those significantly different following multiple comparison correction, and yellow shaded regions are significant prior to correction. DTI- diffusion tensor imaging; FA- fractional anisotropy; DKI- diffusion kurtosis imaging; MK- mean kurtosis; RK- radial kurtosis; NODDI- neurite orientation dispersion and density imaging; SM- standard model; ODI- orientation dispersion index; *p*_2_- orientational coherence; NDI- neurite density index; *f*- intra neurite signal fraction; RISH- rotationally invariant spherical harmonics; R0- rotationally invariant signal for order=0, b=6000 s/mm^2^; D_eperp_- extra axonal perpendicular diffusivity.

#### AOIFCD without head tremor vs. controls

Statistically significant differences were observed in the anterior thalamic radiations (Figure 2), right sided thalamopremotor tract (Figure 3), and left sided striatopremotor tract (Figure 4). Bilateral anterior thalamic radiations showed lower FA (p=1.32×10^-3^) lower RK (p=4.54×10^-4^), higher ODI (p=2.93×10^-4^), coupled with corresponding lower *p*_2_ (p=6.43×10^-4^) and *f* (p=1.63×10^-3^) (standard model) in the dystonia cohort compared to controls. In the right sided thalamopremotor tract higher distal tract ODI (p=8.87×10^-4^), lower *f* (p=3.54×10^-3^) and lower R0 (p=2.26×10^-3^), and mid-tract higher MK (p=1.35×10^-3^), and lower NDI (p=1.65×10^-3^) were observed in the dystonia cohort compared to controls. In the proximal right sided striatopremotor tract lower RO (p=2.94×10^-3^) was observed, with lower mid tract D_eperp_ (p=3.46×10^-3^) on the left side. In the optic tract, the only difference was seen on the right side only in a single measure ( higher ODI, p=1.89×10^-3^) and a single segment (Supplementary Figure 4)

#### AOIFCD with head tremor vs. controls

Significant differences were only observed with NODDI analysis of the frontopontine tract (Supplementary Figure 4), with two regions of lower ODI compared to controls (p=2.4×10^-3^ and p=2.57×10^-3^).

#### Correlation of clinical and imaging parameters

Figure 5 shows the correlation plots for the clinical parameters compared to the significant imaging feature. Significant imaging findings correlated extensively with psychiatric symptom measures across domains.

**Figure 5:**
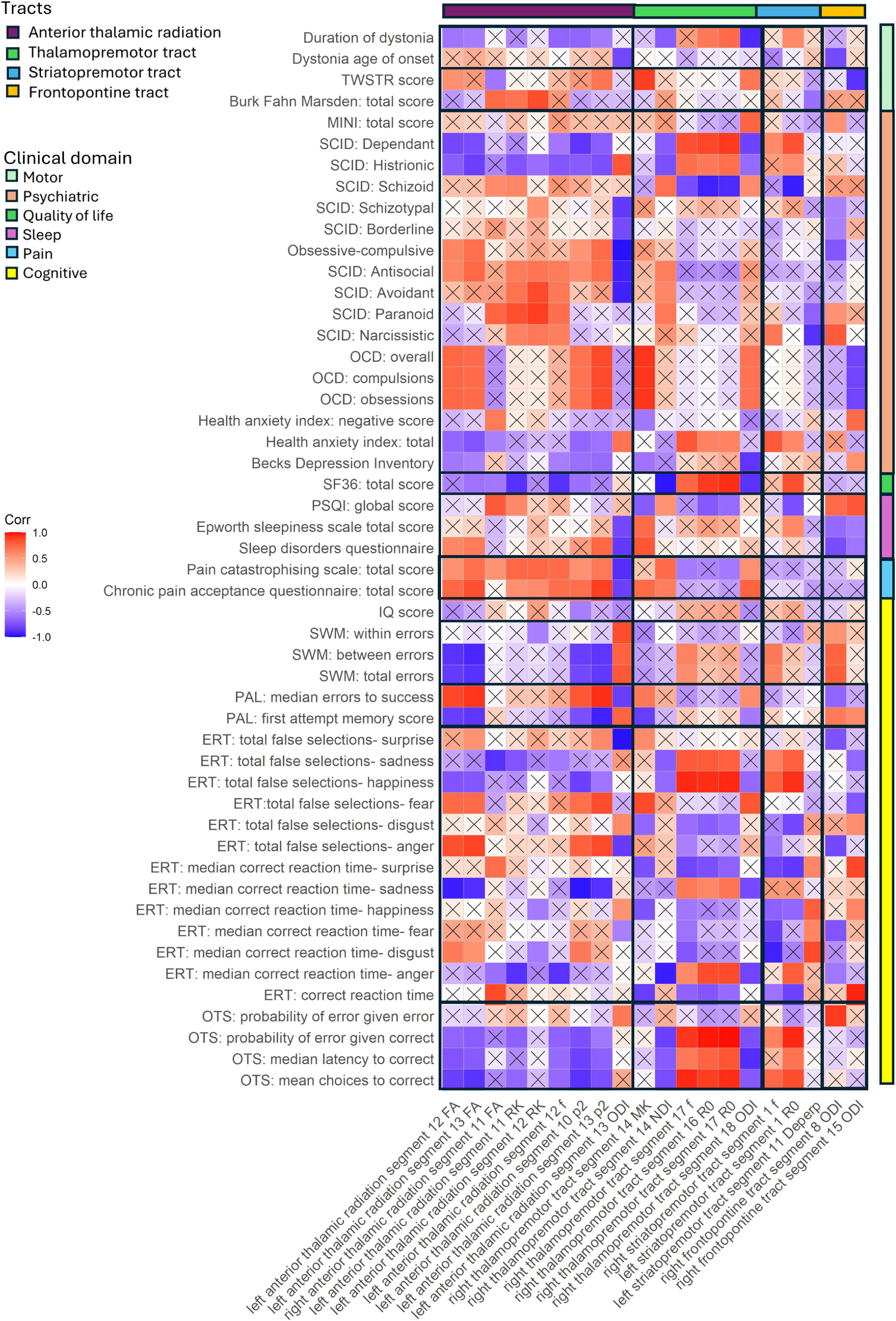
Summary of Pearsons correlation analyses within the dystonia cohort for clinical characteristics and imaging features which were significantly different in dystonia compared to controls. Correlations marked with x were not statistically significant. TWSTR- Toronto West Spasmodic Torticollis Rating scale; MINI- modified mini international neuropsychiatric interview; SCID- Structured clinical interview for DSM-V personality disorders questionnaire; OCD- Yale-Brown Obsessive Compulsive scale; SF36- Short Form 36 Health survey; PSQI- Pittsburgh Sleep Quality Index; SWM- spatial working memory; PAL- paired associated learning; ERT- emotional recognition task; OTS- one touch stockings of Cambridge; FA- fractional anisotropy; MK- mean kurtosis; RK- radial kurtosis; ODI- orientation dispersion index; *p*_2_- orientational coherence; NDI- neurite density index; *f*- intra neurite signal fraction; R0- rotationally invariant signal for L=0, b=6000 s/mm^2^; D_eperp_- extra axonal perpendicular diffusivity.

The left anterior thalamic radiation mid-tract RK correlated with Burke Fahn Marsden score (r=0.85). ODI negatively correlated with SCID personality traits including obsessive-compulsive traits (r=- 0.97), and positively correlated with OCD scores (r=0.92). The SF36 quality of life questionnaire showed anterior thalamic radiation correlations across metrics, the strongest as a negative correlation with mid tract *f*,( r=-0.88). Sleep symptoms correlated with ODI (sleep disorders questionnaire total score: r=-0.84). Widespread tract correlations were seen with pain scores, with the strongest a negative correlation with ODI (r=-0.89). Amongst the CANTAB cognitive testing, correlations were seen across measures in the Spatial Working Memory task (Anterior thalamic radiation mid tract FA, r=-0.91), Paired associated learning task (anterior thalamic radiation mid tract p2, r=0.94), emotional recognition task (sadness; anterior thalamic radiation mid tract p2, r=-0.96).

The right thalamopremotor tract distal changes were negatively correlated with duration of dystonia (ODI, r=-0.9) and mid tract changes positively correlated with TWSTR score (MK, r=0.9). There was correlation with schizoid (r=-0.93) and dependent (r=0.91) personality traits, quality of life (mid tract negative correlation with NDI, r=-0.96, distal tract positive correlation with R0, r=0.94), and sleep (sleep disorders questionnaire, MK r=0.82). Amongst the CANTAB cognitive testing, correlations were seen particularly in the emotional recognition task (sadness; *f*, r=0.95) and executive functioning task (distal thalamopremotor tract R0, r=0.99).

The striatopremotor tract measures included correlations in domains including personality traits (Schizoid, R0: r=-0.94) and quality of life (SF36, R0, r=0.84). Cognitive domains were additionally associated, including in the emotional recognition task (happiness, r=0.93) and executive functioning- (r=0.95). The frontopontine tract measures correlated with TWSTR (r=-0.87), obsessive compulsive symptoms (r=0.8), sleep quality (r=-0.81) and in the emotional recognition task (r=0.96).

## Discussion

Examining one of the largest reported AOIFCD cohorts to date, this study identified multiple localised white matter motor pathway differences in the dystonia cohort compared to controls, most notably involving the anterior thalamic radiations, thalamopremotor tracts and the striatopremotor tracts (Figure 6). These differences were not observed when evaluating whole tract averages (tractography) or during tractometry analysis of other motor tracts (middle cerebellar peduncle, bilateral inferior cerebellar peduncles, superior cerebellar peduncles, corticospinal tracts, superior thalamic radiations, Supplementary table 2) or the optic tracts used as a non-motor comparison. Amongst the general/non-specific measures, statistically significant white matter pathway differences were predominantly observed in FA, MK and RK, while measures of orientational dispersion (ODI, *p*_2_) and intra-axonal signal fraction measures (NDI, *f*) were the most discriminative microstructural modelling characteristics.

**Figure 6:**
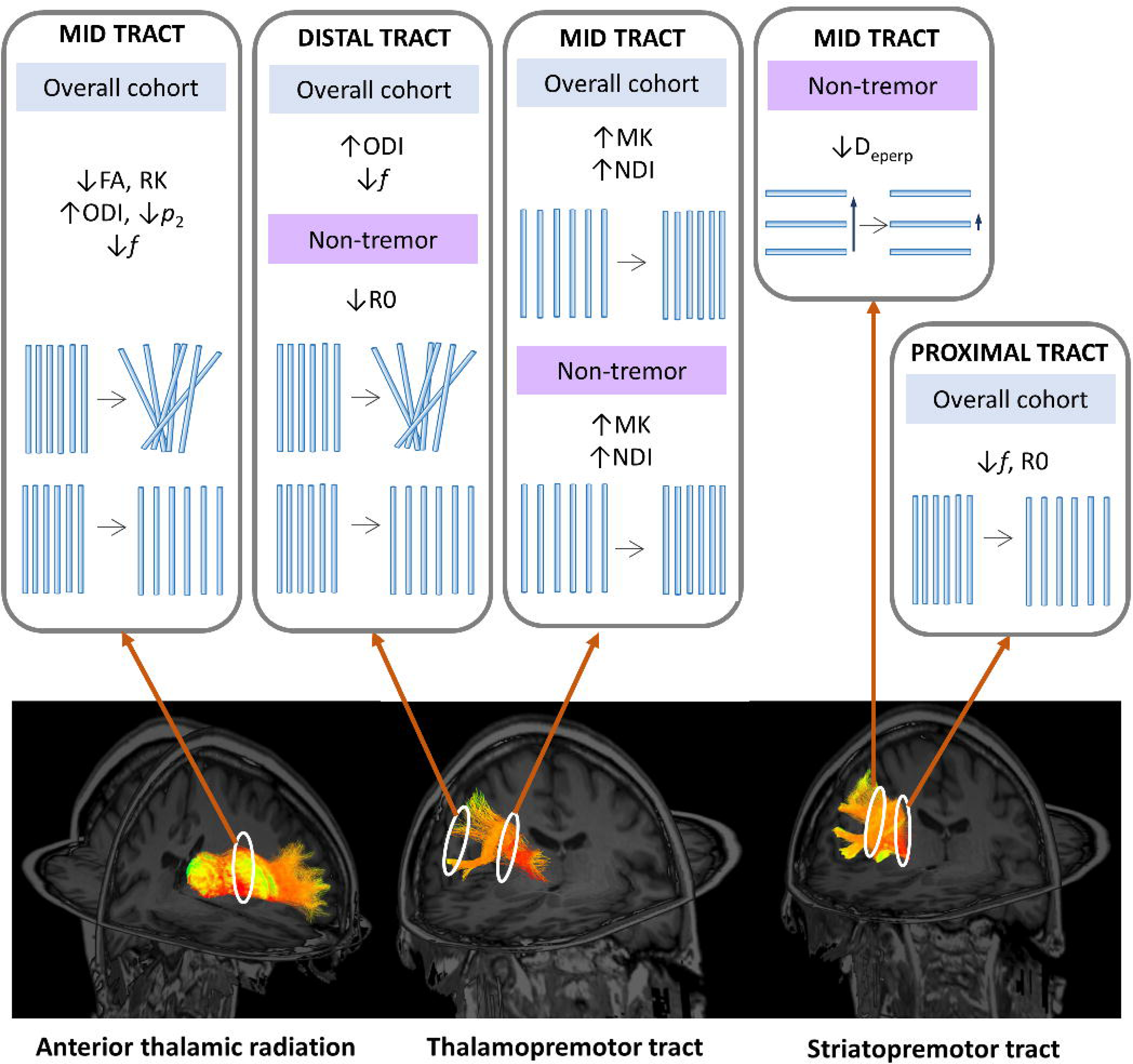
Summary of key findings. FA- fractional anisotropy; MK- mean kurtosis; RK- radial kurtosis; ODI- orientation dispersion index; *p*_2_- orientational coherence; NDI- neurite density index; *f*- intra neurite signal fraction; R0- rotationally invariant signal for L=0, b=6000 s/mm^2^; D_eperp_- extra axonal perpendicular diffusivity.

Differences in AOIFCD seen in the anterior thalamic radiations involved measures associated with lower mid-tract anisotropy (FA), greater orientational dispersion (ODI &*p*_2_) and lower intra axonal signal fraction (*f*), predominantly involving the left sided tracts. By contrast the differences observed in the thalamopremotor tracts involved both mid- and distal-portions, with higher mid-tract neurite density measures (NDI) and an associated trend towards lower orientational dispersion, compared to lower tract density and higher orientational dispersion in the distal tract portion as it approaches the premotor cortex (ODI, *f*). Corresponding patterns were seen in both subgroups (AOIFCD with and without tremor), and although these did not reach statistical significance in the tremor cohort, this is likely due to the relatively small overall size of this subgroup. Within the striatopremotor tracts differences were predominantly seen within the proximal tract where it outflows from the striatum, with lower markers of neurite density (*f*, R0), together with an additional mid tract difference seen in extra-axonal perpendicular diffusivity, the only difference identified in an extra-axonal marker in this study.

As outlined above, few imaging studies in dystonia have made use of approaches which assess more specific microstructural features. However, our findings are consistent with a single study in which fixel-based analysis was used to examine a cervical dystonia cohort. Here they identified reduced apparent fibre density in the vicinity of the striatum, consistent with our findings of lower neurite density markers in the proximal outflow of the striatopremotor tract (16). Our previous work involving NODDI analysis of a mixed dystonia cohort recruited via the UK Biobank, identified changes in the superior cerebellar peduncles and distal anterior thalamic radiations in the cervical dystonia cohort. However, in this instance the work was limited by the level of clinical phenotypic data available with analysis of electronic health records. Despite this, higher thalamopremotor mid-tract FA and regions of lower striatopremotor mid tract neurite density measures were identified in the dystonia cohort, compared to controls, but their significance didn’t survive multiple comparison correction(9). The small and localised nature of the identified tract differences indicate that the pathophysiological changes involved in AOIFCD likely represent subtle, potentially adaptive, microstructural processes, as opposed to widespread pathology. This may be indicative of overutilisation of specific motor pathways and underutilisation of others, creating imbalance and resulting in unopposed excess muscle activation.

Projections relating to the premotor cortex, linking with both the thalamus and striatum, have been implicated in this study. This cortical region is considered to play an important role in motor preparation(37), with overactivity of the premotor cortex having been implicated in dystonia with both Positron Emission Tomography (PET) and fMRI studies (1, 38–41). Thalamic projections, modulating activity of the premotor cortex via the thalamopremotor tract, are predominantly excitatory in nature. The findings from this study of higher mid tract neurite density and lower orientational dispersion potentially indicate overutilisation of this portion of the pathway, while the changes in the distal tract as it approaches the premotor cortex (greater fibre dispersion) potentially suggest disorganisation of this excessive excitatory input. The striatopremotor projections, predominantly providing an excitatory link from the premotor cortex to the striatum, demonstrated differences associated with greater orientational dispersion as the tract approaches the striatum, again potentially indicating disorganisation of the excitatory inputs as they approach their target region. Finally, the anterior thalamic radiations largely provide excitatory projections between the prefrontal cortex and thalamus, with the prefrontal cortex involved in higher cortical functions(42) including executive functioning and other recognised non-motor phenotypes associated with dystonia, such as anxiety and depression(43, 44). Here, the anterior thalamic radiations demonstrated less cohesive mid tract orientation, coupled with positive correlation with multiple non-motor symptom severity, predominantly psychiatric and pain related symptoms (Figure 5).

Whilst these microstructural modelling approaches enable more tissue-feature specific measures, limitations of these as with all current microstructural modelling approaches are that the models are overly simplistic representations of the white matter structure, for example axons are represented as simple ‘sticks’ yet comprise subtler shape variations which are not accounted for in current models. Additionally, the models view white matter structure to consist solely of axons, omitting signal contributions from glial cells, cellular projections and the extracellular matrix, the latter having previously been suggested to be of importance in dystonia pathogenesis(45). This study was also limited by the inability of the method to undertake along tract profiling of those tracts linking the subcortical and primary sensorimotor cortices which were therefore omitted from analysis, despite being key potential pathways in dystonia pathogenesis.

Overall, this work identifies localised changes along white matter motor pathways in a deeply phenotyped cohort of patients diagnosed with AOIFCD, compared to controls. White matter projections linking to the premotor and prefrontal cortices appear to be predominantly implicated, with differences in AOIFCD appearing to predominantly impact the dispersion of fibre orientation and intra-axonal compartment measures. Further work is needed to replicate these findings in larger cohorts and coupled with need for histopathological correlation, to gain greater confidence of the underlying structural changes that give rise to the observed imaging differences.

## Data Availability

All data produced in the present study are available upon reasonable request to the authors

## Acknowledgements

We would like to thank the participants who volunteered for this study, and Dystonia UK for their help with recruitment.

This work was supported by an ABN Clinical Research Training Fellowship funded by the Guarantors of Brain (520286) and a Wellcome Trust Translation of Concept Scheme (Institutional Translational Partnership Award) (520958).CMWT was supported by a Veni grant (17331) from the Dutch Research Council (NWO) and the Wellcome Trust [215944/Z/19/Z]. KJP is funded by an MRC Clinician-Scientist Fellowship & Transition Award (MR/P008593/1, MR/V036084/1).The data were acquired at the UK *National Facility for In Vivo MR Imaging of Human Tissue Microstructure* funded by the EPSRC (grant EP/M029778/1), and The Wolfson Foundation, and supported by a Wellcome Trust Investigator Award (096646/Z/11/Z) and a Wellcome Trust Strategic Award (104943/Z/14/Z).

For the purpose of open access, the author has applied a CC BY public copyright licence to any Author Accepted Manuscript version arising from this submission.

---

Supplementary Figure 1: Whole tract median tractography results for diffusion tensor and diffusion kurtosis measures. FA- fractional anisotropy; MD- mean diffusivity, MK- mean kurtosis; AK- axial kurtosis; RK- radial kurtosis.

Supplementary Figure 2: Whole tract median tractography results for NODDI (Neurite Orientation Dispersion and Density Imaging) and for the rotationally invariant spherical harmonic order=0, b=6000). ODI- orientation distribution index, NDI- neurite density index, FWF- free water fraction, R0- rotationally invariant signal for order=0, b=6000).

Supplementary Figure 3: Whole tract median tractography results for the Standard Model parameters. *f*- intra axonal signal fraction, D_a_- intra axonal diffusivity, D_epar_- extra axonal parallel diffusivity, D_eperp_- extra axonal perpendicular diffusivity, *p*_2_- orientational coherence.

Supplementary Figure 4: Along tract profiles for the frontopontine tracts and optic radiations, showing only parameters which showed significant differences in any analyses, with median tract value and standard error for each cohort. Green shaded regions represent those significantly different following multiple comparison correction, and yellow shaded regions are significant prior to correction. DTI- diffusion tensor imaging; FA- fractional anisotropy; DKI- diffusion kurtosis imaging; MK- mean kurtosis; RK- radial kurtosis; NODDI- neurite orientation dispersion and density imaging; SM- standard model; ODI- orientation dispersion index; *p*_2_- orientational coherence; NDI- neurite density index; *f*- intra neurite signal fraction; RISH- rotationally invariant spherical harmonics; R0- rotationally invariant signal for order=0, b=6000 s/mm^2^; D_eperp_- extra axonal perpendicular diffusivity.

Supplementary Table 1: Summary of participant non-motor clinical phenotyping.

Supplementary Table 2: Tractometry results, using regression modelling with age, gender and handedness as co-variates. Corrected significant results are in bold and indicated with **, uncorrected significant results are indicated with *.

## Notes

### Competing Interest Statement

The authors have declared no competing interest.

### Author Declarations

Participant recruitment was via the Welsh movement disorders research network, (Research Ethics Committee reference: 14/WA/0017 (Integrated research application system) IRAS ID: 146495), with clinical phenotyping performed under the Global Myoclonus Dystonia Registry and Non-Motor Symptoms Study (Research Ethics Committee reference: 18/WM/0031, IRAS (Integrated research application system) project ID: 236219). Ethical approval for the brain imaging was obtained via the Cardiff University School of Medicine Research Ethics Committee (SMREC reference number 18/30)

