## Supplementary figures and images for "White matter microstructural changes in adult-onset idiopathic focal cervical dystonia using ultra-strong diffusion gradient MRI"

### Supplementary Figure 1

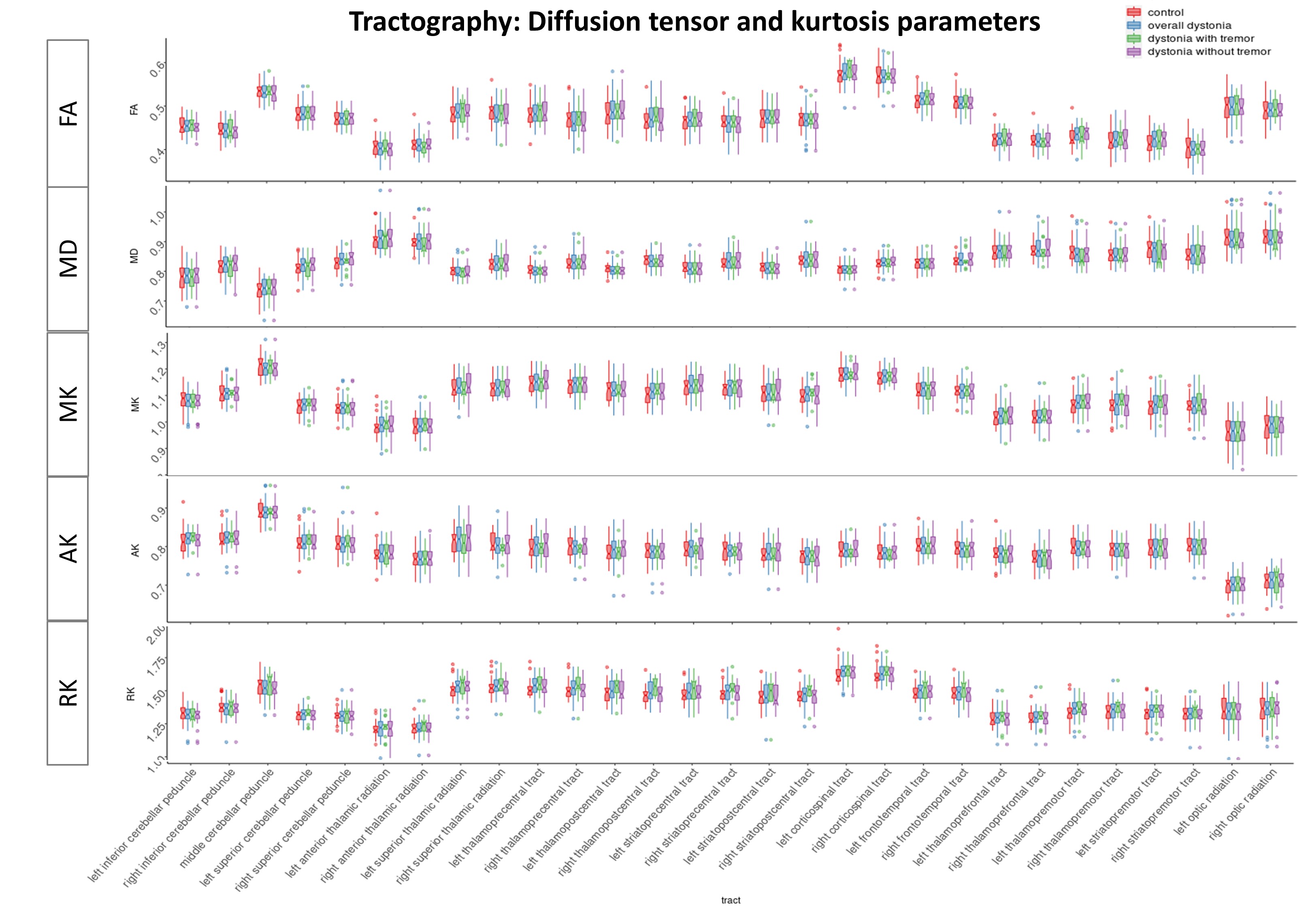

### Supplementary Figure 2

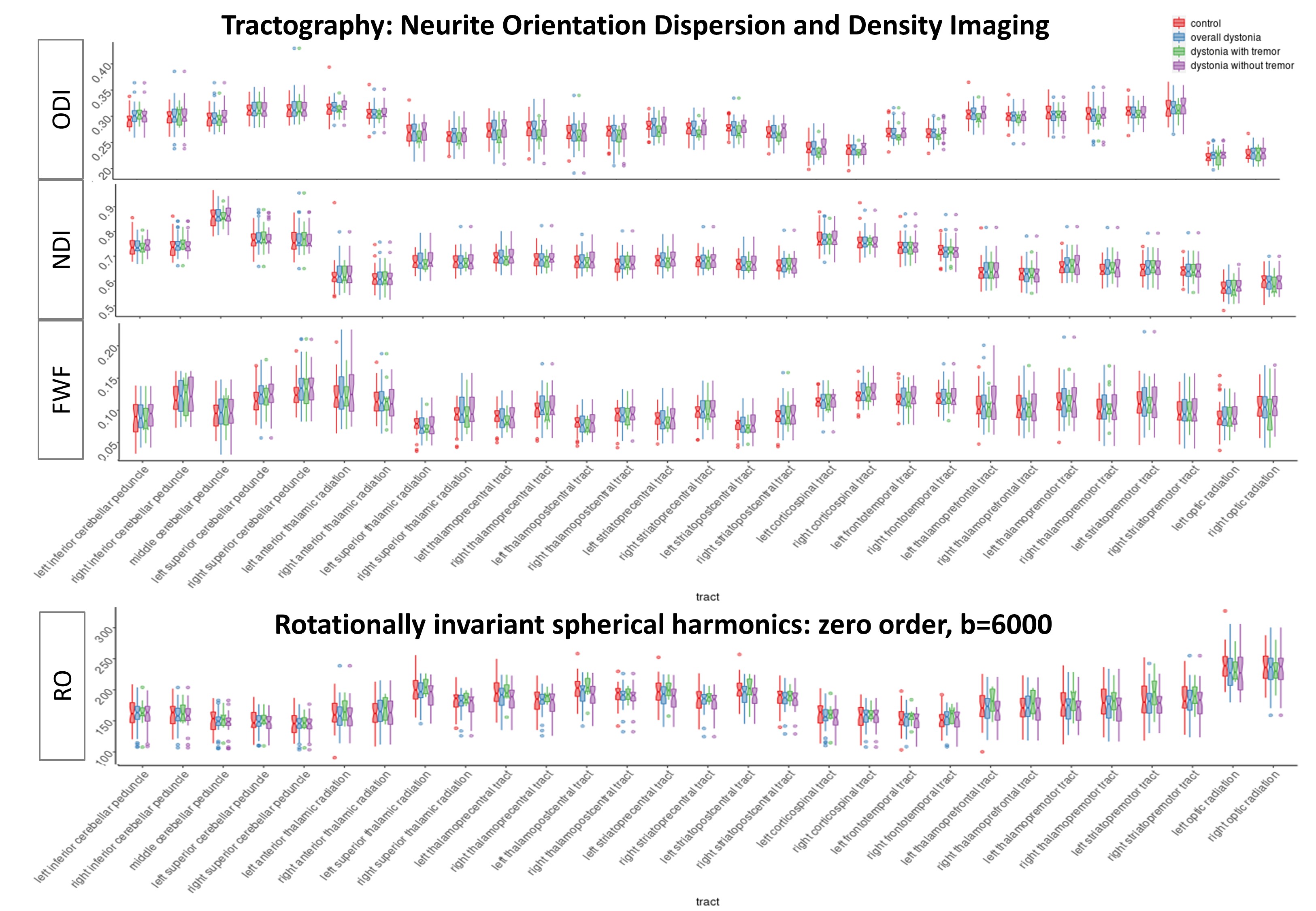

### Supplementary Figure 3

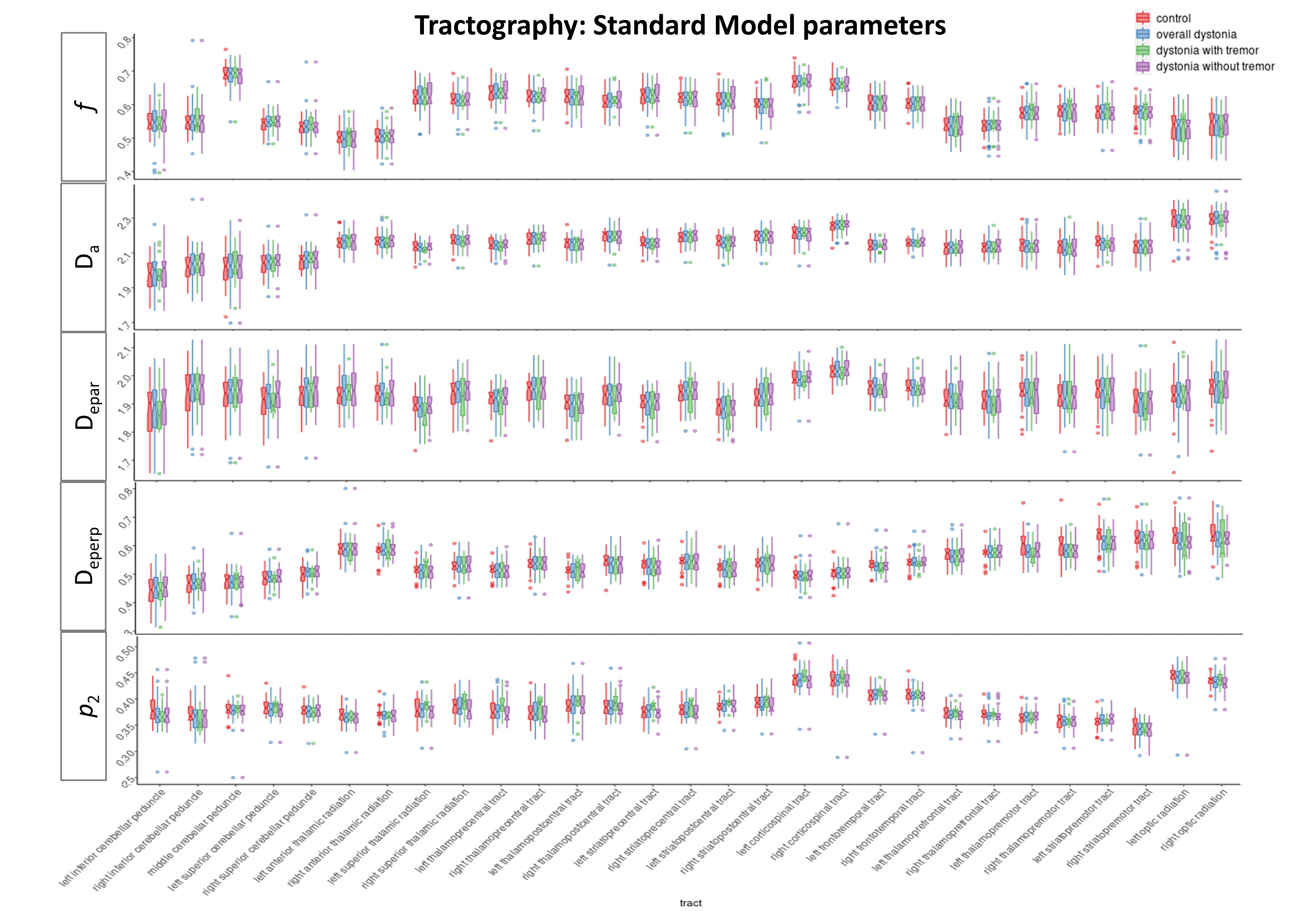

### Supplementary Figure 4

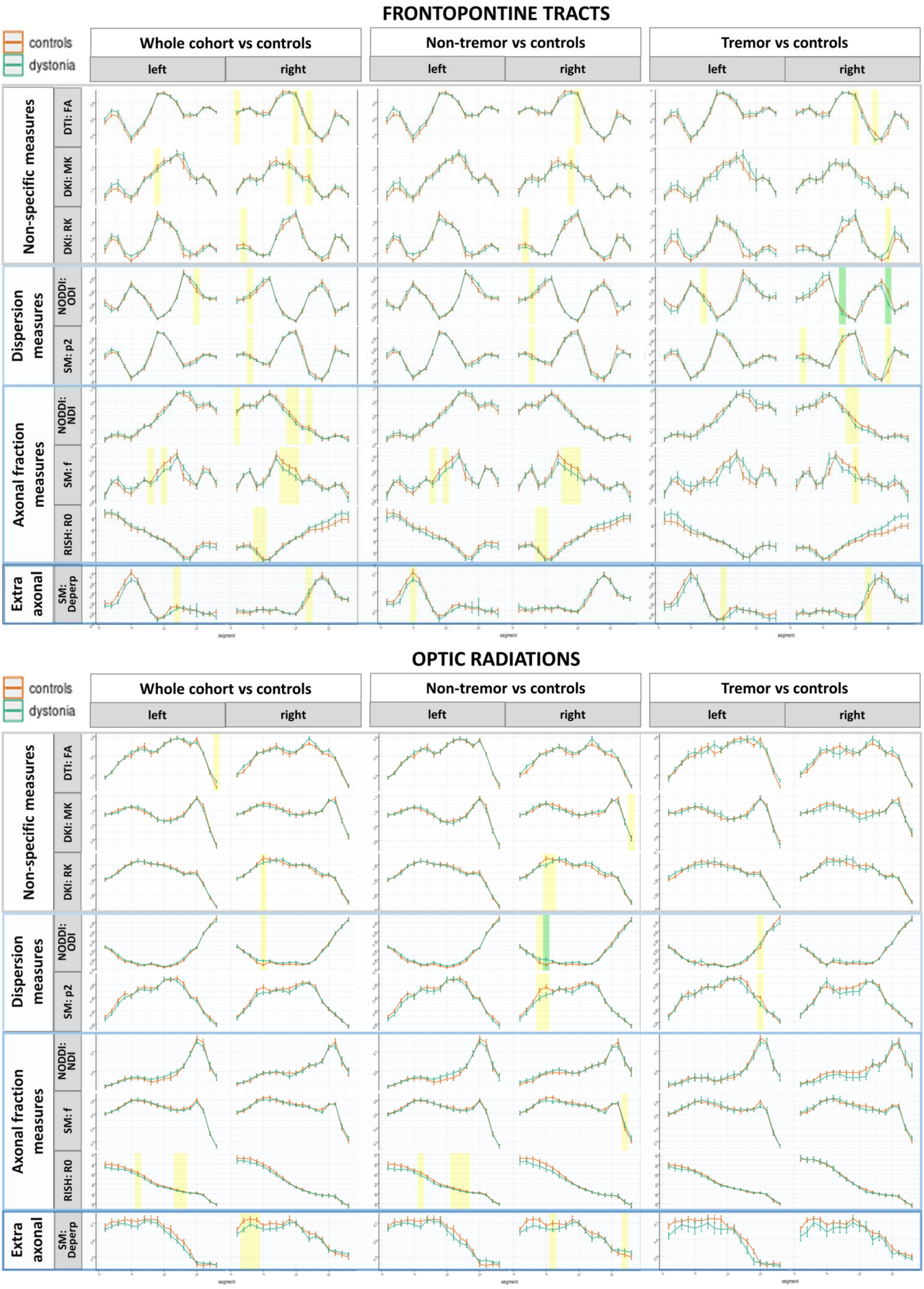
