## Supplementary Table 1 for "White matter microstructural changes in adult-onset idiopathic focal cervical dystonia using ultra-strong diffusion gradient MRI"

| ***Characteristic*** | | | | ***Control cohort*** | | ***Overall AOIFCD*** | | | **AOIFCD without tremor** | | | | | ***AOIFCD with tremor*** | | |
| --- | --- | --- | --- | --- | --- | --- | --- | --- | --- | --- | --- | --- | --- | --- | --- | --- |
|  |  |  |  |  |  | ***Median (range)*** | | ***p- value vs controls*** | ***Median (range)*** | | ***p- value vs controls*** | | | ***Median (range)*** | | ***p-value vs controls*** |
| **Modified MINI Scale** | | | | | | | | | | | | | | | | |
| *Median score (range)* | | | | 1 (0-9) | | 4 (0-15) | **2.49x10^-4^*** | | 4 (0-15) | **5.23x10^-4^*** | | | | 4.5 (0-12) | **0.014*** | |
| *Total score ≥6 (percentage)* | | | | 16 (36%) | | 11 (85%) | **5.38x10^-14^*** | | 11 (85%) | **5.02x10^-13^*** | | | | 5(15%) | **4.23x10^-9^*** | |
| **SCID-5-PD** | | | | | | | | | | | | | | | | |
| Group A | | *Paranoid* | | 1 (0-4) | 1 (0-4) | | 0.545 | | 1 (0-4) | | | 0.6 | 1 (0-4) | | 0.633 | |
|  |  | *Schizoid* | | 1 (0-4) | 2 (0-7) | | 0.197 | | 2 (0-7) | | | 0.227 | 2 (0-2) | | 0.36 | |
|  |  | *Schizotypal* | | 1 (0-5) | 1 (0-7) | | 0.629 | | 1 (0-7) | | | 0.77 | 1 (0-6) | | 0.522 | |
| Group B | | *Antisocial* | | 0 (0-3) | 0 (0-4) | | 0.197 | | 0 (0-2) | | | 0.505 | 1 (0-4) | | **0.048*** | |
|  |  | *Borderline* | | 1 (0-5) | 1 (0-6) | | 0.234 | | 1 (0-6) | | | 0.253 | 2 (0-4) | | 0.451 | |
|  |  | *Histrionic* | | 1 (0-3) | 1 (0-5) | | 0.271 | | 1 (0-5) | | | 0.24 | 1 (0-3) | | 0.643 | |
|  |  | *Narcissistic* | | 1 (0-7) | 1 (0-6) | | 0.012 | | 1 (0-6) | | | 0.031 | 0.5 (0-4) | | **0.04*** | |
| Group C | | *Avoidant* | | 2 (0-7) | 3 (0-8) | | 0.612 | | 2.5 (0-8) | | | 0.838 | 3.5 (0-8) | | 0.384 | |
|  |  | *Dependent* | | 1 (0-3) | 1 (0-5) | | 0.826 | | 1 (0-5) | | | 0.886 | 0.5 (0-4) | | 0.798 | |
|  |  | *Obsessive-compulsive* | | 2 (0-6) | 2 (0-7) | | 0.521 | | 2 (0-6) | | | 0.359 | 2 (0-7) | | 0.861 | |
| **Becks Depression Inventory** | | | | | | | | | | | | | | | | |
| Total score | | | | 8 (1-38) | | 11 (0-38) | **0.0199*** | | 7 (1-14) | **0.032*** | | | | 8 (0-38) | 0.094 | |
| **Health Anxiety index** | | | | | | | | | | | | | | | | |
| Health anxiety inventory | | | | 5.5 (0-17) | | 7 (0-30) | 0.154 | | 8 (0-26) | 0.054 | | | | 6 (1-30) | 0.814 | |
| Negative consequences subscale | | | | 2 (0-8) | | 3 (0-9) | 0.089 | | 3.5 (1-9) | **0.015*** | | | | 1.5 (0-8) | 0.632 | |
| **Yale Brown OCD scale** | | | | | | | | | | | | | | | | |
| Obsessions | | | | 0 (0-10) | | 0 (0-14) | 0.549 | | 0 (0-14) | 0.766 | | | | 0 (0-9) | 0.366 | |
| Compulsions | | | | 0 (0-15) | | 0 (0-13) | 0.85 | | 0 (0-13) | 0.883 | | | | 0 (0-13) | 0.865 | |
| Global severity sore | | | | 0 (0-25) | | 0 (0-27) | 0.944 | | 0 (0-27) | 0.941 | | | | 0 (0-22) | 0.988 | |
| **Sleep related domains** | | | | | | | | | | | | | | | | |
| *Pittsburgh sleep quality index* | *Subjective sleep quality* | | | 1 (0-3) | | 1.5 (0-3) | **0.019*** | | 1 (0-3) | **0.038*** | | | | 2 (0-3) | **0.0551*** | |
|  | *Sleep latency* | | | 1 (0-3) | | 1 (0-3) | **0.013*** | | 1 (0-3) | **0.013*** | | | | 1 (0-3) | 0.179 | |
|  | *Sleep duration* | | | 1 (0-2) | | 1 (0-3) | 0.567 | | 1 (0-3) | 0.9 | | | | 1 (0-3) | 0.078 | |
|  | *Habitual sleep efficiency* | | | 0.5 (0-3) | | 1 (0-3) | 0.096 | | 1 (0-3) | 0.124 | | | | 1 (0-3) | 0.233 | |
|  | *Sleep disturbances* | | | 2 (2-3) | | 3 (1-3) | 0.1 | | 3 (2-3) | **0.033*** | | | | 2 (1-3) | 0.837 | |
|  | *Use of sleeping medication* | | | 0 (0-2) | | 0 (0-3) | 0.21 | | 0 (0-3) | 0.418 | | | | 0 (0-3) | 0.086 | |
|  | *Daytime dysfunction* | | | 1 (0-2) | | 1 (0-3) | **0.001*** | | 1 (0-3) | **0.006*** | | | | 1 (0-2) | **0.008*** | |
|  | *Global score* | | | 6.5 (2-15) | | 9 (2-20) | **0.004*** | | 9 (3-20) | **0.007*** | | | | 8.5 (2-14) | 0.052 | |
| *Sleep Disorders Questionnaire* | *Total score* | | | 9 (6-26) | | 18 (7-50) | **5.78x10^-5^*** | | 17 (7-50) | **2.29x10^-4^*** | | | | 20.5 (7-40) | **0.004*** | |
| *Epworth Sleepiness Scale* | *Total score* | | | 5 (0-15) | | 6 (0-20) | 0.308 | | 6 (0-20) | 0.388 | | | | 6 (0-9) | 0.387 | |
| **Pain related domains** | | | | | | | | | | | | | | | | |
| *Presence of pain (number- yes:no, percent)* | | | | 11:18 | | 37:7 | **2.5x10^-4^*** | | 28:4 | **1.7x10^-4^*** | | | | 9:3 | **0.69*** | |
| *Chronic pain acceptance questionnaire* | | | *Activity engagement* | 57 (33-64) | | 39 (3-65) | **6.03x10^-4^*** | | 40 (3-65) | **0.012*** | | | | 34 (12-57) | 0.184 | |
|  |  |  | *Pain willingness* | 20.5(3-35) | | 29 (0-54) | 0.064 | | 29 (0-45) | 0.076 | | | | 26 (14-54) | 0.165 | |
|  |  |  | *Total score* | 59 (35-73) | | 66 (47-94) | 0.089 | | 66 (47-94) | 0.171 | | | | 63 (54-74) | 0.538 | |
| *Pain catastrophising scale* | | | *Rumination* | 5 (4-14) | | 8 (4-20) | **0.048*** | | 8 (4-18) | 0.098 | | | | 12 (4-20) | **0.042*** | |
|  |  |  | *Magnification* | 4 (3-6) | | 5 (3-12) | **0.026*** | | 5 (3-10) | 0.069 | | | | 7 (3-12) | **0.015*** | |
|  |  |  | *Helplessness* | 7 (6-15) | | 12 (6-28) | **0.003*** | | 11 (6-25) | **0.01*** | | | | 15 (7-28) | **0.0052*** | |
|  |  |  | *Total score* | 5 (0-22) | | 12 (0-47) | **0.004*** | | 7 (0-40) | **0.015*** | | | | 22 (2-47) | **0.005*** | |
| **Quality of life** | | | | | | | | | | | | | | | | |
| *Sf-36 Health survey* | | | *Physical functioning* | 90 (35-100) | | 60 (5-100) | **2.31x10^-4^*** | | 60 (15-100) | **4.79x10^-4^*** | | | | 67.5 (5-100) | **0.014*** | |
|  |  |  | *Physical role limitation* | 100 (0-100) | | 12.5 (0-100) | **2.69x10^-4^*** | | 25 (0-100) | **0.001*** | | | | 0 (0-100) | **0.003*** | |
|  |  |  | *Bodily pain* | 73.75 (42.5-80) | | 55 (30-80) | **1.03x10^-4^*** | | 55 (30-80) | **2.91x10^-4^*** | | | | 42.5 (30-80) | **0.004*** | |
|  |  |  | *General health perceptions* | 75 (40-95) | | 55 (10-95) | **3.3x10^-4^*** | | 57.5 (10-85) | **1.88x10^-4^*** | | | | 55 (10-95) | 0.104 | |
|  |  |  | *Energy/vitality* | 60 (45-80) | | 55 (40-70) | **0.014*** | | 55 (40-70) | **0.042*** | | | | 50 (40-60) | **0.02*** | |
|  |  |  | *Social functioning* | 100 (12.5-100) | | 62.5 (0-100) | **3.9x10^-5^*** | | 68.75 (0-100) | **1.6x10^-4^*** | | | | 37.5 (0-100) | **0.002*** | |
|  |  |  | *Emotional role limitations* | 100 (33.33-100) | | 100 (0-100) | **0.002*** | | 100 (0-100) | **0.01*** | | | | 50 (0-100) | **0.0013*** | |
|  |  |  | *Mental health* | 68 (56-84) | | 64 (40-84) | 0.162 | | 66 (40-84) | 0.244 | | | | 64 (44-76) | 0.182 | |
| **CANTAB: One Touch Stockings of Cambridge (executive functioning task)** | | | | | | | | | | | | | | | | |
| *Mean choices to correct (OTSMCC)* | | | | 1.365 (1-3.6) | | 1.5 (1-3.6) | **0.03*** | | 1.47 (1-2.87) | 0.066 | | | | 1.635 (1.07-3.6) | 0.065 | |
| *Median latency to correct (OTSMDLC)* | | | | 14256 (5927-50269) | | 17831.5 (5213-53102) | 0.161 | | 15431 (5213-48102) | 0.5 | | | | 22030.5 (7461-53102) | **0.034*** | |
| *Probability of error given error (OTSPEGE)* | | | | 0.2 (0-0.83) | | 0.33 (0-1) | **0.002*** | | 0.29 (0-1) | **0.016*** | | | | 0.44 (0-0.92) | **0.002*** | |
| *Probability of error given correct (OTSPEGC)* | | | | 0.285 (0-1) | | 0.4 (0-1) | 0.13 | | 0.365 (0-1) | 0.326 | | | | 0.47 (0.07-1) | 0.069 | |
| **CANTAB: Emotional Recognition task** | | | | | | | | | | | | | | | | |
| Median correct reaction time (ERTCRT) | | | | 1164.5 (762-2534.5) | | 1399.5 (719.5-3478) | 0.054 | | 1399.5 (719.5-2170) | 0.091 | | | | 1357.25 (945-3478) | 0.129 | |
| Median correct reaction time: anger (ERTCRTA) | | | | 1528 (667.5-4695) | | 1645 (778-4128) | 0.674 | | 1678.25 (778-4062) | 0.84 | | | | 1645 (912-4128) | 0.523 | |
| Median correct reaction time: disgust (ERTCRTD) | | | | 1550.5 (753.5-3767.5) | | 1420 (803.5-3913) | 0.845 | | 1320 (803.5-3495) | 0.554 | | | | 2070.25 (828-3913) | 0.123 | |
| Median correct reaction time: fear (ERTCRTF) | | | | 1979 (976-4595) | | 2119 (1092-7421) | 0.319 | | 2087 (1092-7421) | 0.427 | | | | 2278 (1219.5-3429) | 0.361 | |
| Median correct reaction time: happiness (ERTCRTH) | | | | 968.5 (611-2528) | | 958 (553-2695.5) | 0.758 | | 903 (553-1875.5) | 0.77 | | | | 1026 (728-2695.5) | 0.212 | |
| Median correct reaction time: Sadness (ERTCRTS) | | | | 1461.5 (636.5-3895.5) | | 1545 (695-4328.5) | 0.49 | | 1510.5 (695-3346) | 0.683 | | | | 1817 (809-4328.5) | 0.371 | |
| Median correct reaction time: surprise (ERTCRTSU) | | | | 1095 (623-2329) | | 1370.25 (461-3879) | **0.02*** | | 1311.75 (461-2912) | 0.086 | | | | 1454 (970-3879) | **0.015*** | |
| Total false selections: anger (ERTTFAA) | | | | 2 (0-10) | | 1 (0-8) | **0.008*** | | 0 (0-8) | **0.002*** | | | | 1 (0-6) | 0.512 | |
| Total false selections: disgust (ERTTFAD) | | | | 4 (0-15) | | 3.5 (0-12) | 0.78 | | 3.5 (0-12) | 0.782 | | | | 3.5 (2-8) | 0.878 | |
| Total false selections: fear (ERTTFAF) | | | | 2 (0-7) | | 3 (0-8) | 0.687 | | 2 (0-8) | 0.842 | | | | 3.5 (0-7) | 0.185 | |
| Total false selections: happiness (ERTTFAH) | | | | 1.5 (0-11) | | 2 (0-12) | 0.107 | | 2 (0-12) | 0.17 | | | | 2.5 (1-4) | 0.169 | |
| Total false selections: sadness (ERTTFAS) | | | | 3 (0-11) | | 4 (0-12) | 0.431 | | 3 (0-12) | 0.842 | | | | 5 (0-10) | 0.134 | |
| Total false selections: surprise (ERTTFASU) | | | | 5 (0-12) | | 4 (0-11) | 0.582 | | 4 (0-11) | 0.395 | | | | 5.5 (0-11) | 0.839 | |
| **CANTAB: Paired Associated Learning** | | | | | | | | | | | | | | | | |
| *First attempt memory score (PALFAMS)* | | | | 15 (5-20) | | 12 (2-20) | **0.034*** | | 14 (2-20) | 0.332 | | | | 8.5 (3-16) | **0.001*** | |
| *Mean errors to success (PALMETS)* | | | | 2 (0-4) | | 1 (0-6) | 0.15 | | 1 (0-6) | 0.204 | | | | 1.5 (0-3) | 0.262 | |
| *Number of patterns reached (PALNPR)* | | | | 8 (6-8) | | 8 (4-8) | **0.014*** | | 8 (4-8) | 0.2 | | | | 6 (4-8) | **1.58x10^-4^*** | |
| **CANTAB: Spatial Working Memory** | | | | | | | | | | | | | | | | |
| *Total errors (SWMTE)* | | | | 6.5 (0-26) | | 16.5 (0-36) | **0.003*** | | 16 (0-30) | **0.012*** | | | | 18.5 (1-36) | **0.011*** | |
| *Between errors (SWMBE)* | | | | 6 (0-26) | | 16.5 (0-34) | **0.003*** | | 16 (0-28) | **0.014*** | | | | 18.5 (1-34) | **0.012*** | |
| *Within errors (SWMWE)* | | | | 0 (0-4) | | 0 (0-13) | **0.034*** | | 0 (0-13) | 0.051 | | | | 0 (0-9) | 0.074 | |
| *Between and within errors (SWMDE)* | | | | 0 (0-2) | | 0 (0-7) | 0.088 | | 0 (0-7) | 0.168 | | | | 0 (0-7) | 0.077 | |

*Supplementary table 1: Clinical phenotyping assessment results. Significant differences are bold and indicated with *. AOIFCD- adult- onset idiopathic focal cervical dystonia; MINI- Mini International Neuropsychiatric Interview; SCID-5-PD- Structured clinical interview for DSM 5 personality disorders; OCD- obsessive compulsive disorder; CANTAB- Cambridge Neuropsychological Test Automated Battery.*
